# Combined diffusion-weighted MRI and diffusion-weighted MR spectroscopy for interrogating the glioma tumour microenvironment: a preliminary study

**DOI:** 10.64898/2026.07.23.26356578

**Authors:** Marco Palombo, Matteo Figini, Samuel Rot, Elizabeth Powell, Bhavana Solanky, Chloé Najac, Bernard Siow, Jeremy Rees, Eleftheria Panagiotaki, Anestis Passalis, Claudia Wheeler-Kingshott, Itamar Ronen, Harpreet Hyare

## Abstract

**Background and purpose:** Gliomas are characterised by a complex tumour microenvironment (TME) that contributes to treatment resistance and tumour heterogeneity. Therefore, the non-invasive interrogation of both the intracellular and extracellular compartments of gliomas remains a key unmet need. We investigated the feasibility and complementarity of combining diffusion-weighted MRI (DW-MRI) with biophysical modelling Vascular, Extracellular, and Restricted Diffusion for Cytometry in Tumours (VERDICT) and diffusion-weighted MR spectroscopy (DW-MRS) for simultaneous characterisation of glioma tumour cells and the glioma TME.

**Methods:** 14 patients with newly diagnosed glioma (WHO grades 2–4: 4 IDH-wildtype and 10 IDH-mutant) underwent DW-MRI at 3 T; DW-MRS was additionally acquired in 10 patients. Tumours were automatically segmented into enhancing, non-enhancing, and oedema regions using a validated pipeline. VERDICT models were fitted to multi-shell DW-MRI data to estimate intracellular volume fraction (f*IC*), cell radius, extracellular diffusivities (D and D), and free-water fraction (f*FW*). Single voxel DW-MRS (20×20×20 mm³) provided metabolite-specific apparent diffusion coefficients (ADCs) for total N-acetylaspartate (tNAA), Creatine (tCr), and choline (tCho). T-tests assessed DW-MRI and descriptive statistics assessed DW-MRS parameters in tumour regions compared to normal appearing white matter (NAWM) and IDH mutation status. Pearson’s correlations assessed associations between DW-MRS metabolite ADCs and DW-MRI parameters.

**Results:** VERDICT-MRI distinguished high grade IDH-wildtype from lower grade IDH-mutant gliomas, with significantly higher f*IC* and lower D and D in enhancing and non-enhancing regions of IDH-wildtype lesions. DW-MRS demonstrated a trend towards reduction in tNAA ADC in tumour versus contralateral NAWM, consistent with neuronal loss, and a trend towards increased tCho ADC, suggesting glial activation. A descriptive trend towards decreased tNAA ADC in IDH-wildtype tumours was observed. Significant positive correlations were identified between tumoral tCho ADC and VERDICT parameters f*EES*, D and negative correlations for ADC and f*FW*, in non-enhancing tumour regions.

**Conclusion:** This proof-of-concept study demonstrates the feasibility of combining multi b-value DW-MRI and DW-MRS within a clinically feasible protocol to simultaneously probe the extracellular and intracellular compartments of the glioma TME. VERDICT captured cell-level and extracellular matrix differences in IDH mutation status, while DW-MRS provided metabolite-specific indices of neuronal and glial compartment integrity. The correlation between tCho ADC and VERDICT metrics in infiltrative tumour regions supports the complementarity of these modalities. With this combined approach, it is possible to simultaneously characterise the tumour compartment and the tumour microenvironment in gliomas.

## 1. Introduction

Glioma is the most common primary brain tumour in adults, with the most aggressive form glioblastoma multiforme (GBM) characterised by rapid proliferation, diffuse infiltration, and near-universal recurrence despite multimodal therapy comprising of surgery, radiotherapy, and temozolomide chemotherapy. (Incekara et al., 2020; Prager et al., 2020) Treatment resistance arises from multiple converging mechanisms, including tumour heterogeneity, immune evasion, and the profound influence of the tumour microenvironment (TME). (Prager et al., 2020)

The glioma TME encompasses a complex ecosystem of cellular and non-cellular constituents: endothelial cells, neurons, astrocytes, oligodendrocytes, resident microglia, tumour-associated macrophages, tumour-infiltrating lymphocytes, and non-cellular components including paracrine signalling molecules, exosomes, and extracellular matrix (ECM) components. (Martinez-Lage et al., 2019) The TME plays a vital role in promoting tumour cell survival and conferring resistance to therapy via complex bidirectional signalling networks that govern biomass synthesis, metabolic reprogramming, and immune suppression. (Plaks et al., 2015; Hanahan and Coussens, 2012; Frisch et al., 2019) Mapping these dynamic TME interactions non-invasively, in addition to tumour cell profiling is therefore paramount for understanding tumour biology and for developing more effective therapeutic strategies.

Diffusion-weighted (DW) MRI exploits sensitivity to the microscale displacement of water molecules to probe tissue microstructure in vivo (Basser and Pierpaoli, 2011; Wedeen et al., 2012; Norris, 2001). Advanced biophysical models of the DW-MRI signal, such as Neurite Orientation Dispersion and Density Imaging (NODDI) (Zhang et al., 2012) and Vascular, Extracellular, and Restricted Diffusion for Cytometry in Tumours (VERDICT) (Panagiotaki et al., 2014; Panagiotaki et al., 2015), decompose the signal into compartment-specific contributions, providing quantitative indices of neurite density, cell size, intracellular volume fraction, free water fraction and extracellular diffusivity. Although developed to characterize the brain tissue, NODDI has been increasingly used to characterize the TME, with increasing evidence of added value for tumour grading and assessment of treatment response (Maximov et al., 2017; Zerweck et al., 2025; Zhao et al., 2025). VERDICT was initially validated for solid tumour characterisation (Panagiotaki et al., 2014; Panagiotaki et al., 2015) and has recently been adapted for brain tumours, demonstrating the ability to capture cellular and vascular microstructural changes in glioma (Roberts et al., 2020; Figini et al., 2023).

DW-MRS leverages the fact that brain metabolites are almost exclusively confined to the intracellular space, offering a unique non-invasive window on cell-specific compartments without relying on assumptions about the extracellular water signal (Ligneul et al., 2024; Palombo et al., 2018). *N*-acetylaspartate (NAA) and glutamate are preferentially localised in neurons (Gill et al., 1989; Simmons et al., 1991; Petroff et al., 1993); myo-inositol (Ins) and total choline (tCho) are enriched in glial cells, particularly astrocytes (Choi et al., 2007); and total creatine (tCr) is relatively evenly distributed across cellular compartments. By measuring the ADC of these metabolites, DW-MRS can therefore directly and non-invasively characterise the diffusion environment experienced within cells of distinct types (Palombo et al., 2018; Palombo et al., 2016). Consensus recommendations for DW-MRS acquisition and analysis have now been established (Ligneul et al., 2024).

By combining biophysical modelling of DW-MRI (VERDICT and NODDI) with DW-MRS, it is in principle possible to interrogate both the extracellular and intracellular compartments of gliomas simultaneously, providing complementary microstructural and metabolic information that neither technique can deliver alone. However, the feasibility of this combined approach in glioma patients has not been systematically evaluated. In this preliminary study, we applied both modalities in a 45 minute protocol to a cohort of 14 patients with newly diagnosed glioma spanning a range of WHO grades and IDH mutation status. The aim was to assess whether (i) the combined approach is technically and clinically feasible and (ii) the two modalities provide concordant and complementary biological information. Our hypothesis is that the cell-type specificity of DW-MRS and DW-MRI can not only capture information from the tumor cells but also the tumour microenvironment and help disambiguate the interpretation of water-based DW-MRI.

## 2. Methods

### 2.1 Patients

Fourteen patients (6 male, mean age 44.1 ± 17.2 years, range 22–76 years) with newly diagnosed glioma were prospectively recruited and imaged following written informed consent, in accordance with local ethics approval (05/Q0502/101 (34772-PHYSICS01)). Eleven patients were imaged pre-operatively and three were imaged following surgical resection but with residual disease (patients 5, 7 and 11). Histopathological diagnoses comprised: WHO grade 4 IDH-wildtype GBM (n = 4), WHO grade 3 IDH-mutant astrocytoma (n = 1) and WHO grade 2 IDH-mutant astrocytoma and oligodendroglioma (n = 9). Full clinical and conventional imaging characteristics are provided in Table 1.

**Table 1.**
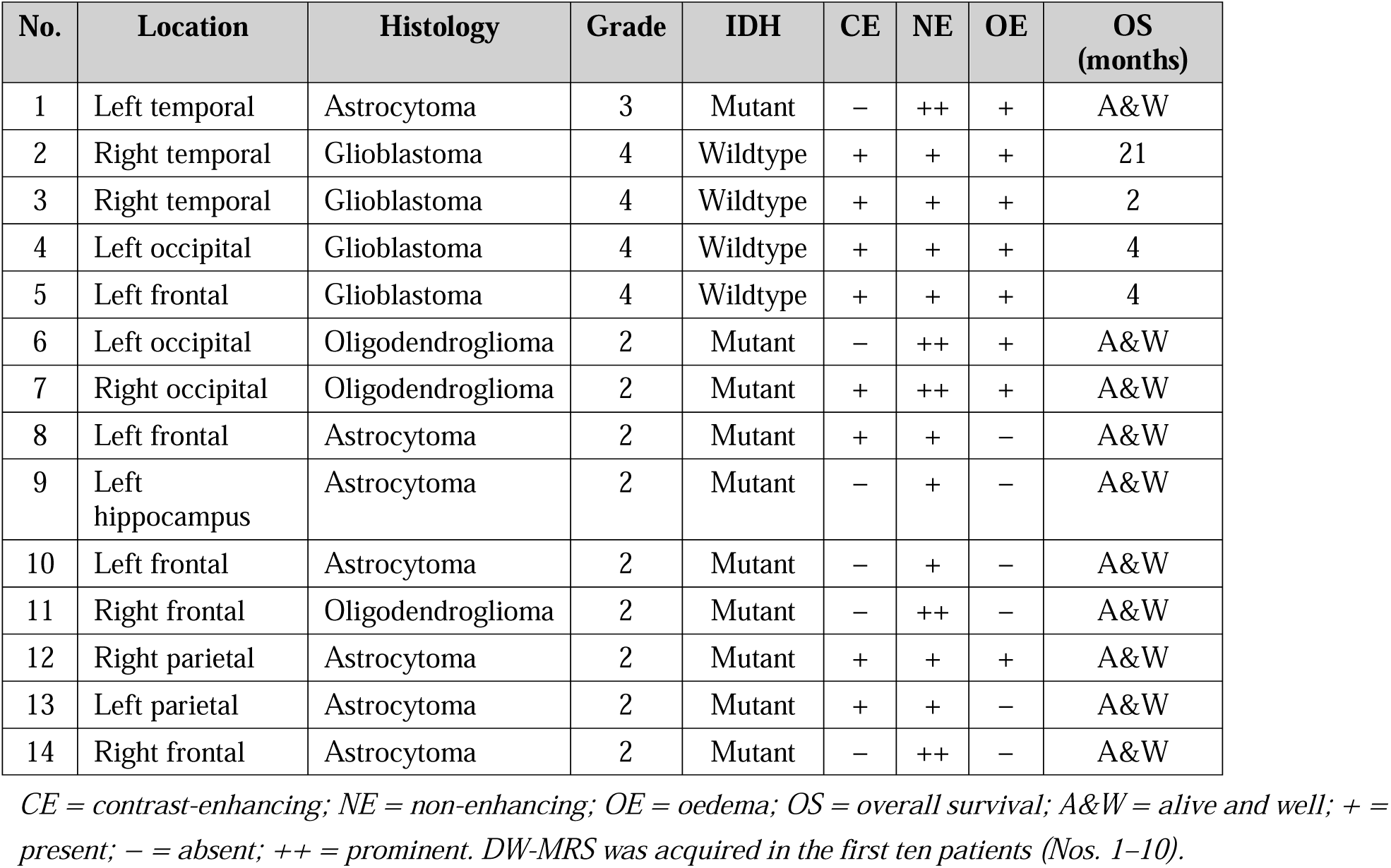
Clinical and conventional imaging characteristics of recruited patients.

### 2.2 DW-MRI acquisition

All MRI was performed on a 3 T clinical scanner (Ingenia CX, Philips Healthcare, Best, Netherlands). DW-MRI was acquired with 2 mm isotropic resolution and comprised 10 shells with 3 diffusion-encoding directions and variable b-values (50–3500 s/mm²), gradient pulse durations δ (5–20 ms), and separation times Δ (25–95 ms) (see supplementary Table 1), plus 2 additional shells with 24 diffusion directions at b = 711 s/mm² and 48 directions at b = 2500 s/mm², δ = 24 ms, Δ = 45 ms. Total scan time for the diffusion protocol was approximately 35 minutes. Three-dimensional T1-weighted and fluid-attenuated inversion recovery (FLAIR) images were also acquired for anatomical reference and segmentation.

### 2.3 DW-MRS acquisition

DW-MRS was acquired in the first ten patients in addition to DW-MRI. The DW-MRS sequence (Nilsson et al., 2018) was ported to the clinical Philips Ingenia CX system and optimised to: (a) reduce peripheral nerve stimulation; and (b) prevent inadvertent rotation of diffusion gradients with the spectroscopy voxel. A voxel of interest (VOI) 20 × 20 × 20 mm³ was positioned in the tumour core (according to consultant neuroradiologist) and in contralateral normal appearing white matter (NAWM) using a T1-weighted scan for planning. A water-suppressed PRESS sequence was used with a bipolar diffusion-weighting scheme; cardiac triggering (PPU peripheral pulse unit) was applied to achieve an effective TR of 3 RR intervals. Other acquisition parameters: TE = 90 ms, spectral bandwidth = 2000 Hz, 1024 data points, 108 dynamic scans. Gradient amplitudes of 5, 20, and 40 mT/m were applied along three orthogonal directions, yielding b-values of 60, 956, and 3823 s/mm², respectively. An unsuppressed water reference acquisition was performed for eddy current correction.

### 2.4 DW-MRI analysis

DW-MRI data were denoised (Veraart et al., 2016) and corrected for Gibbs ringing artefacts (Kellner et al., 2016), susceptibility distortions (Andersson and Sotiropoulos, 2016), and motion (Nilsson et al., 2018) prior to modelling. The NODDI model was fitted to the last two shells (b = 711 and b = 2500 s/mm²) using the NODDI toolbox (https://www.nitrc.org/projects/noddi_toolbox) to estimate neurite orientation dispersion index (ODI) and intra-neurite volume fraction (f*ICVF*). The free-water fraction (f*ISO*) estimated by NODDI was used to constrain the free-water fraction (f*FW*) of the four-compartment VERDICT model (Figini et al., 2023). VERDICT was then fitted to the full multi-shell DW-MRI dataset using in-house MATLAB code to estimate: intracellular signal fraction (f*IC*), cell radius (*R*), extracellular/extravascular signal fraction (f*EES*), axial and radial extracellular diffusivities (D and D), and vascular signal fraction (f*VASC*) (Figini *et al*., 2023). The standard apparent diffusion coefficient (ADC) was computed from b = 1000 s/mm² data as a reference metric.

### 2.5 DW-MRS analysis

Spectral registration across transients was applied within each diffusion-weighting condition to correct for phase and frequency drift, and transients exhibiting a marked drop in signal amplitude due to non-translational motion were discarded prior to combination (n=2). Spectral quantification was performed with LCModel using an appropriate basis set. Estimates for tNAA (NAA + NAAG), tCr (Cr + PCr), and tCho (Cho + PCho + GPC) were extracted and used to compute metabolite-specific ADCs from the three b-values. Spectra with a Cramér–Rao lower bound > 20% for the metabolite of interest, or exhibiting poor spectral quality on visual inspection, were excluded from analysis; this resulted in missing values for some patients, particularly within the tumour voxel: valid tNAA data: 3/8 tumour, 7/8 NAWM; valid tCr and tCho data: 4/8 tumour, 7/8 NAWM.

### 2.6 Tumour segmentation

Tumours were segmented using an established in-house automated pipeline (Ruffle et al., 2023; Ruffle et al., 2024) incorporating automated sequence identification, parameter determination, and image pre-processing (denoising, registration, super-resolution). Tumours were segmented into BraTS-standard regions: *1. enhancing tumour*, *2. non-enhancing tumour*, and *3. peri-lesional oedema* (Figure 1), following the approach described by Ruffle *et al*. (Ruffle et al., 2023; Ruffle et al., 2024). Segmentations were quality-controlled and edited where necessary by a neuroradiologist with 15 years of clinical experience. NAWM regions of interest (ROIs) were manually drawn in contralateral normal-appearing white matter for each patient.

**Figure 1:**
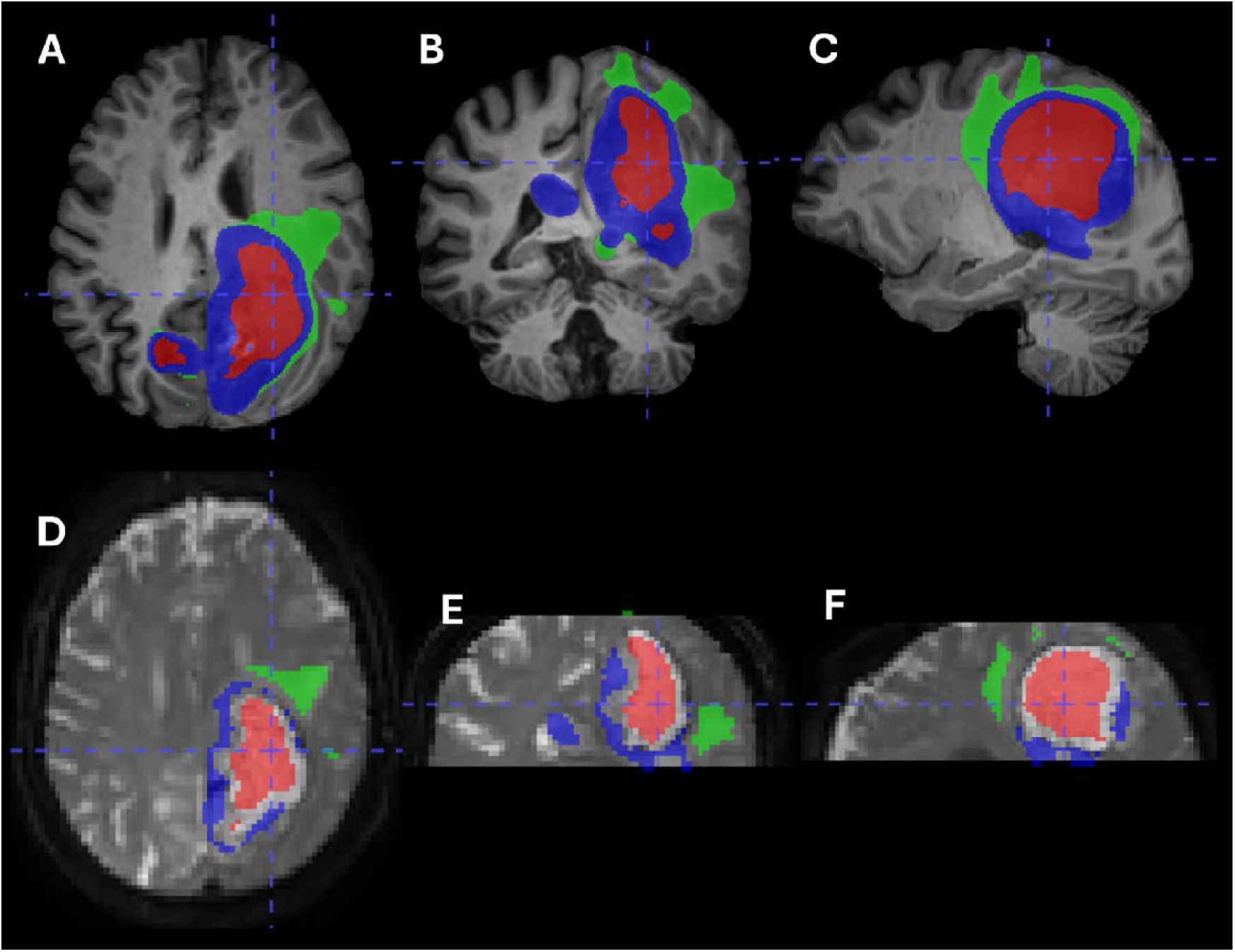
Representative tumour segmentations according to BraTS criteria. IDH-wildtype left occipital GBM (case n.4). A–C: segmented tumou components overlaid onto the T1-weighted MRI are illustrated as follows: red = enhancing; green = non-enhancing; blue = oedema. D–F illustrate the tumour segmentations applied to the b = 0 volume of the DW-MRI data following linear registration of T1-weighted images.

To register tumour ROIs to the space of the DW-MRI–derived parameter maps, T1-weighted images were linearly registered to the b = 0 volume of the DW-MRI data using the FLIRT algorithm in FSL. The estimated transformations were then applied to the binarised ROI masks.

### 2.7 Statistical analysis

Median values of VERDICT and ADC metrics were extracted per region per patient. Group differences between IDH-wildtype and IDH-mutant lesions were assessed using two-sample t-tests, with p < 0.05 considered statistically significant. Pearson’s correlation coefficients were used to assess associations between DW-MRS metabolite ADCs and DW-MRI parameters in corresponding tumour and NAWM regions. Due to small sample sizes, Mann-Whitney U test and descriptive statistics were used to assess differences in tumour versus NAWM VOI and IDH-mutant versus IDH-wildtype tumours. All statistical analyses were performed in MATLAB.

## 3. Results

### 3.1 DW-MRI

Figure 2 shows representative T1-weighted images, ADC maps, and VERDICT parameter maps for a WHO grade IV IDH-wildtype GBM and a WHO grade II IDH-mutant astrocytoma. As expected, the GBM exhibits higher f*IC* (indicative of elevated cellularity) and greater spatial heterogeneity across all maps compared with the lower-grade lesion.

**Figure 2:**
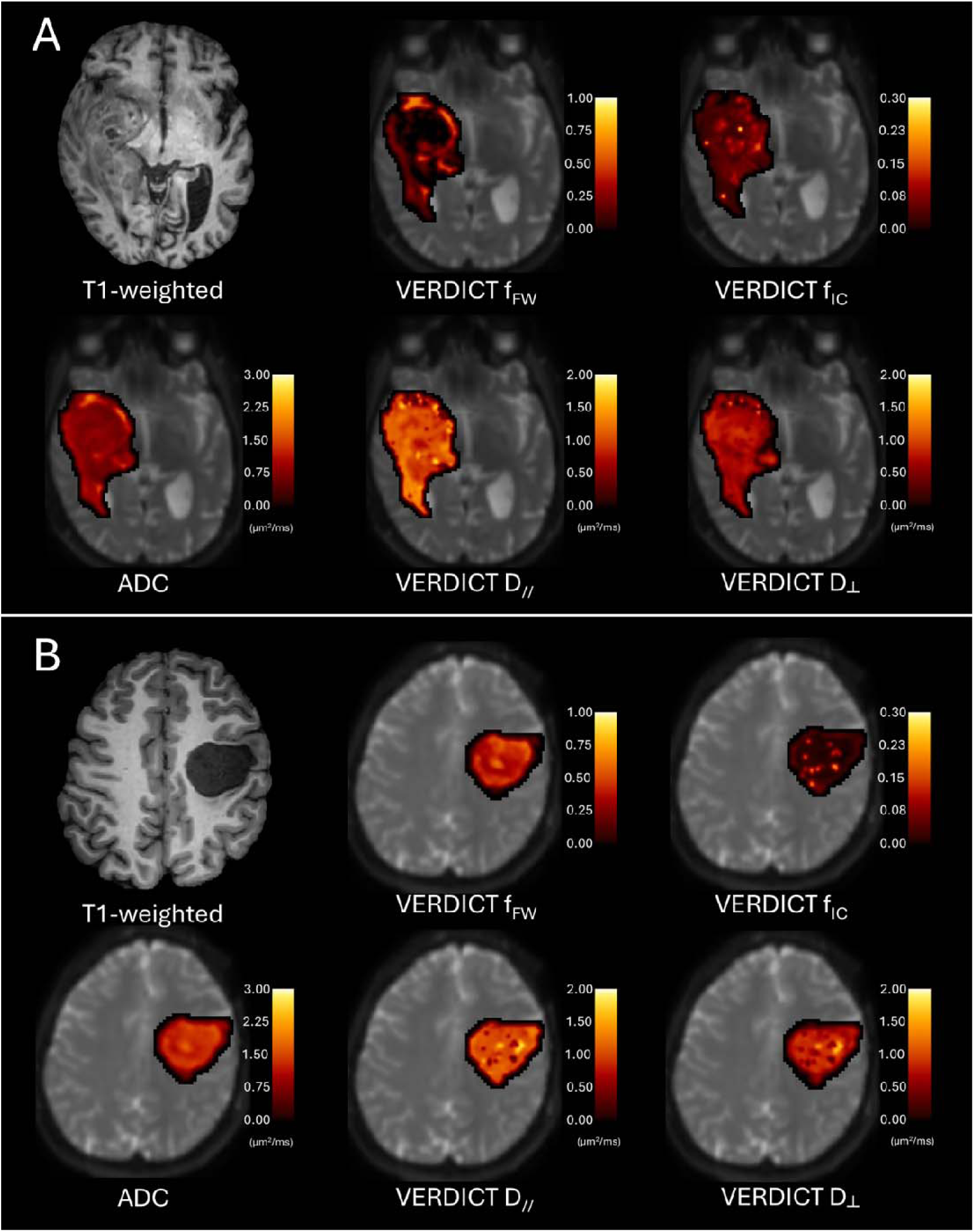
Representative T1-weighted image and ADC and VERDICT maps. A. Right temporal GBM (case n. 3). B. IDH-mutant WHO grade 2 left frontal astrocytoma (case n. 8). ADC and VERDICT maps are displayed in colour within the tumour area and overlaid on the b = 0 DW-MRI volume.

Figure 3 summarises the quantitative DW-MRI group comparison between IDH-wildtype and IDH-mutant lesions. In IDH-wildtype lesions, compared with IDH-mutant lesions, the median ADC was significantly lower in enhancing tumour areas and showed a non-significant trend towards higher values in non-enhancing areas. The free-water fraction f*FW* followed the same pattern as ADC. The intracellular fraction f*IC* was significantly higher in IDH-wildtype enhancing tumour, with no significant difference in other sub-regions. Notably, extracellular diffusivities (D and D) were significantly lower in both enhancing and non-enhancing tumour areas of IDH-wildtype lesions. No statistically significant differences between IDH mutations were observed in oedema regions. NODDI-derived fICVF and ODI were estimated but are not reported as primary outcomes here, as their interpretation is less straightforward in tumour tissue than in normal brain and their role was to constrain the VERDICT free-water fraction.

**Figure 3:**
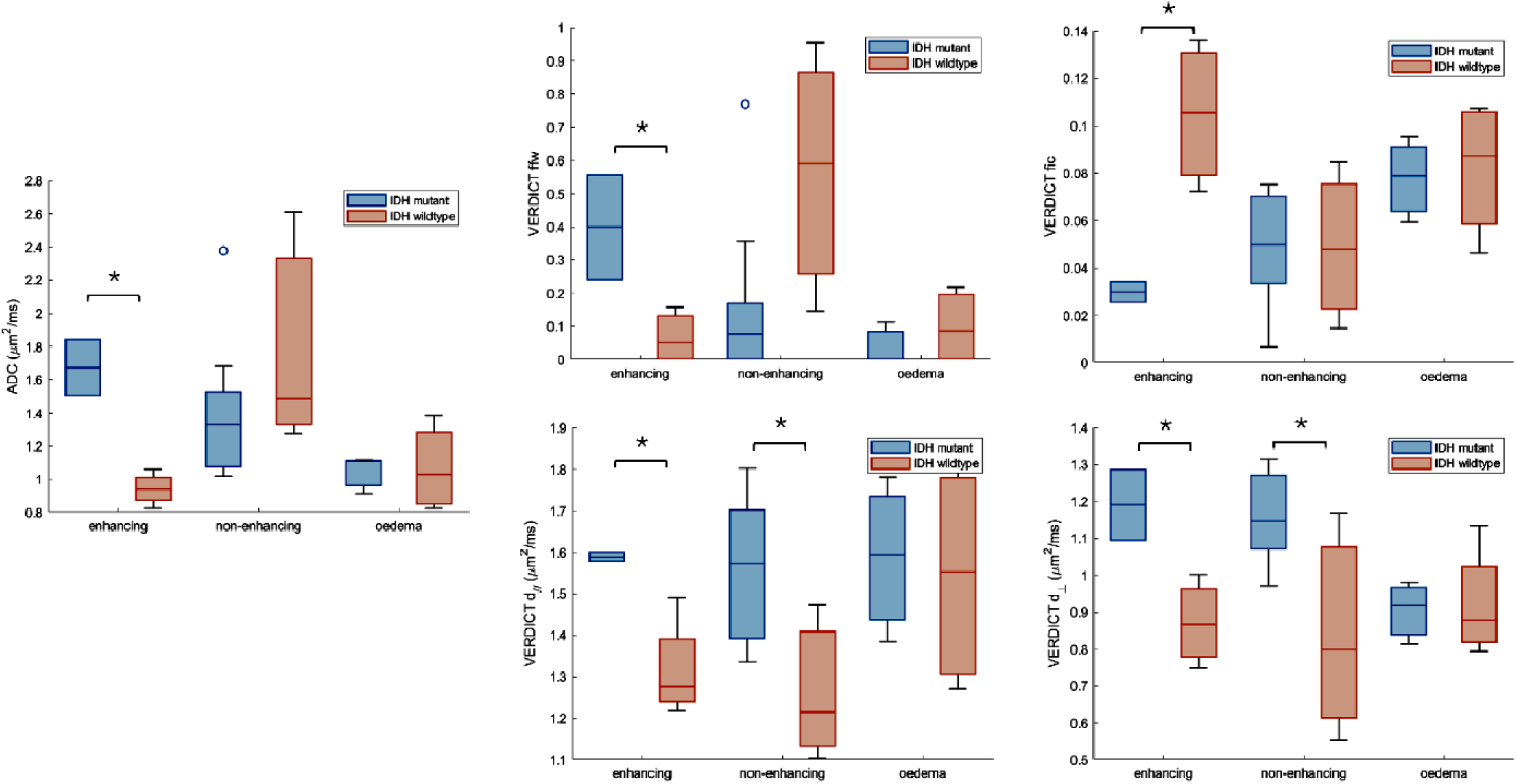
Quantitative DW-MRI analysis in IDH-mutant and IDH-wildtype gliomas. The figure shows boxplots of median ADC and VERDICT metrics (ffw, fic, d_//_ and d^) across patients in the IDH-mutant and IDH-wildtype groups. Statistically significant differences (t-test, p<0.05) are marked by asterisks. VERDICT metric f_FW_ was significantly lower in enhancing areas, and non-significantly higher in non-enhancing areas; f_IC_ was significantly higher in enhancing areas and showed no statistically significant difference in other areas. The extracellular diffusivities (d_//_ and d^) were lower in both enhancing and non-enhancing areas. No statistically significant difference was found in oedema compartment. Tumour components: enhancing: IDH-mut n=2, IDH-wt n=4; non-enhancing: IDH-mut n=9, IDH-wt n=4; oedemas: IDH-mut n=3, IDH-wt n=4

### 3.2 DW-MRS

#### Metabolite ADC in tumour versus NAWM

Descriptive statistics showed a trend towards reduced tNAA ADC in tumour compared with contralateral NAWM consistent with neuronal atrophy or loss. tCho ADC showed a modest, non-significant increase in tumour versus NAWM, which may reflect glial activation or reactive astrocytosis. Neither metabolite reached significance (p=0.38 and p=0.01 respectively), most likely due to being underpowered.

#### Metabolite ADC and IDH mutation status

Although group sizes precluded formal statistical comparison, a descriptive trend of decreased tNAA ADC in IDH-wildtype versus IDH-mutation was observed within tumour VOIs. This suggests that higher-grade gliomas are associated with greater alterations in the neuronal component of the TME. In contrast, tCho ADC appeared stable across IDH mutation status, indicating minimal grade-dependent variation in the glial compartment. Representative DW spectra from contralateral and tumour VOIs are displayed in Figure 4.

**Figure 4:**
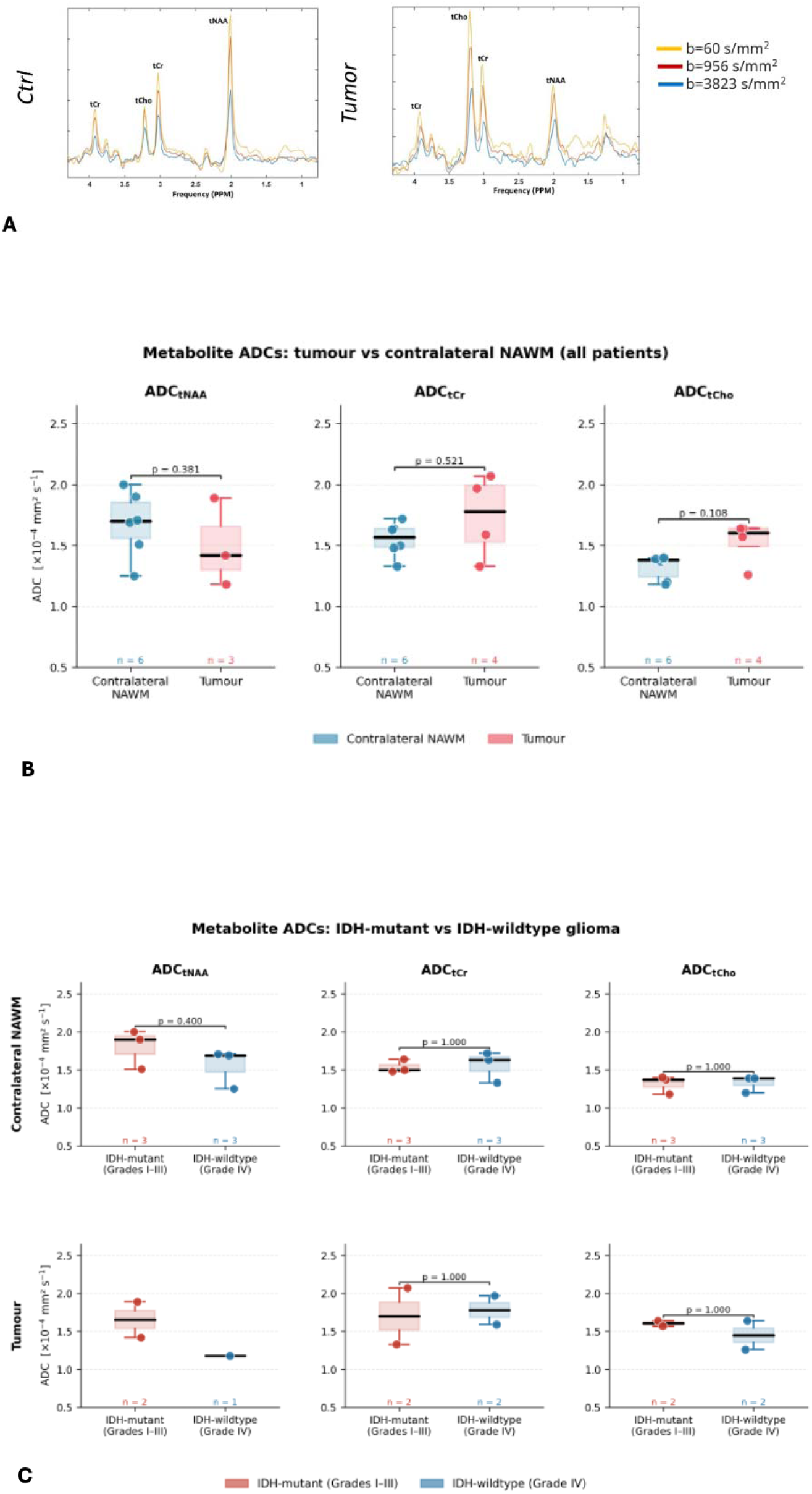
ADC of major brain metabolites. A. Representative spectra in tumour and contralateral NAWM VOI. B. Changes in ADC of major brain metabolites in tumour versus contralateral NAWM VOI. Decrease in tNAA ADC and increase in tCho ADC which did not reach significance was observed in tumour versus NAWM VOI. C reports the metabolite changes with respect to IDH mutation status. Although group sizes precluded formal statistical comparison, a descriptive trend of decreased tNAA ADC in IDH-wildtype versus IDH-mutation was observed within tumour VOIs. tCho ADC was stable across IDH mutation status.

### 3.3 Correlations between DW-MRI and DW-MRS

Significant correlations were found between tumoral ADC*tCho* and the following DW-MRI parameters in non-enhancing tumour ROIs: ADC, f*FW*, f*EES*, and D (Figure 5). No other significant correlations were identified, including in NAWM or for other metabolites. It should be noted that the limited number of valid DW-MRS observations and the slight imbalance in available data across metabolites severely constrain statistical power, particularly for tNAA ADC; no significant correlation survived correction for multiple comparisons.

**Figure 5:**
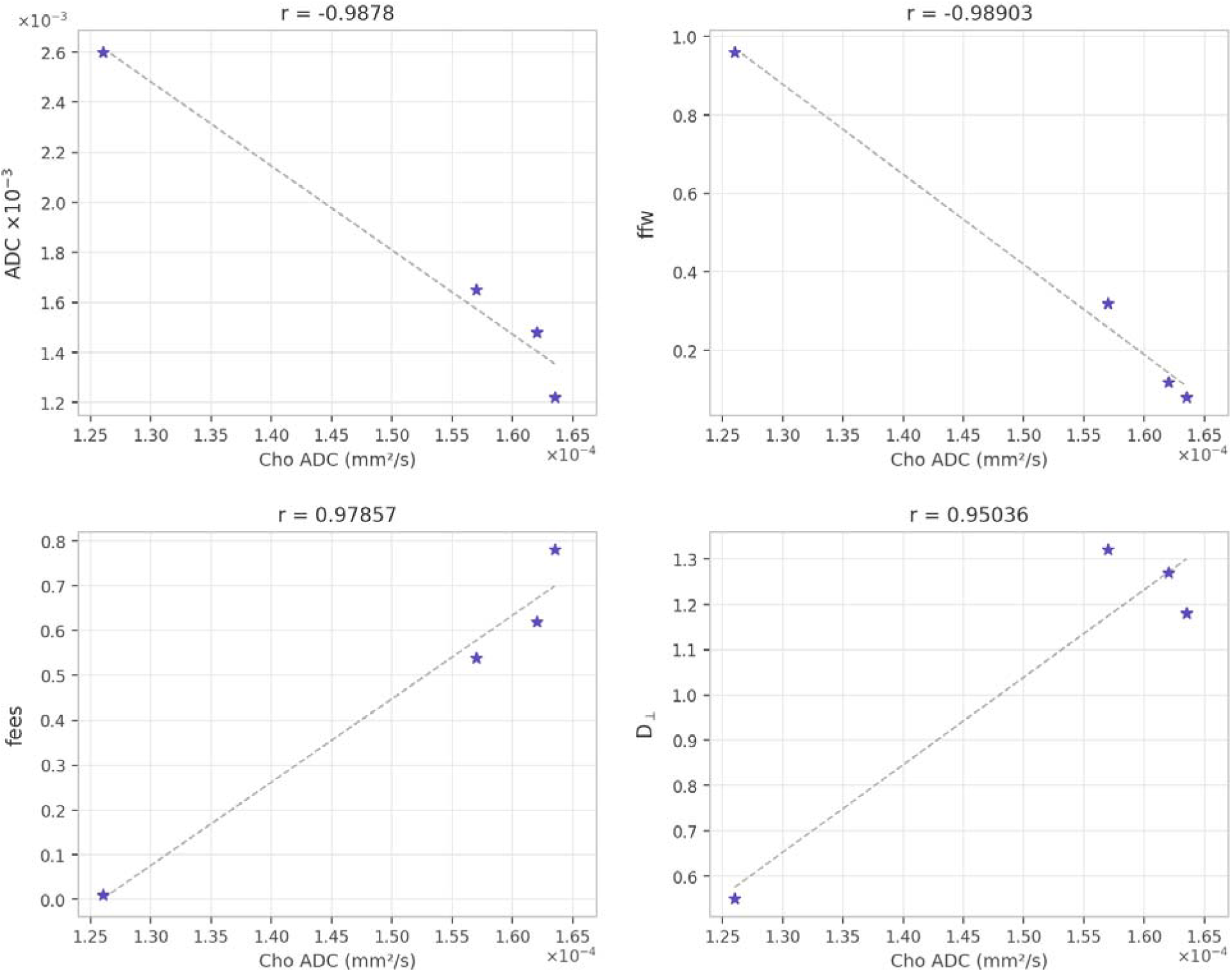
Correlation of DW-MRI and DW-MRS. Significant correlations were found between tumoural tCho ADC and the following DW-MRI parameters in non-enhancing ROIs: ADC, f_FW_, f_ees_ and D^. No significant correlations were observed in the other tumour components (including NAWM) or amongst the other metabolites.

## 4. Discussion

This proof-of-concept study demonstrates the feasibility of combining DW-MRI and DW-MRS for non-invasive characterisation of the glioma TME in a clinical setting. The principal findings are threefold. First, metabolite ADCs measured by DW-MRS were sensitive to tumour-related changes, with tNAA ADC reduced in tumour relative to NAWM, consistent with neuronal atrophy, and tCho ADC modestly elevated, compatible with increase in the glial compartment. Second, VERDICT-derived DW-MRI metrics differentiated high grade IDH-wildtype from lower grade IDH-mutant tumours, with higher f*IC* and lower extracellular diffusivities in the more aggressive IDH-wildtype phenotype, suggesting higher tumour cellularity and increase in extracellular matrix components. Third, tCho ADC correlated with several VERDICT metrics in non-enhancing tumour regions, characterising infiltrative tumour. VERDICT-MRI captured cell-level and extra-cellular matrix (ECM)-level differences between IDH mutation strata, while DW-MRS provided metabolite-specific measures of neuronal and glial compartment integrity. By combining biophysical modelling of DW-MRI (VERDICT) with DW-MRS, we have demonstrated that it is possible to interrogate both the extracellular and intracellular compartments of gliomas simultaneously for improved glioma characterisation.

Our DW-MRS findings leverage previous work distinguishing and quantitatively characterizing brain cell morphologies noninvasively (Palombo et al., 2016; Ronen et al., 2014; De Marco et al., 2022). Prior DW-MRS work in healthy rodent and primate brain using Monte Carlo simulation-based biophysical modelling has consistently supported preferential localisation of Ins and tCho inside astrocytes and Glu and NAA in neurons, with no preferential compartment for tCr (Palombo et al., 2016; Ronen et al., 2014; De Marco et al., 2022; Li et al., 2014; Hall and Alexander, 2009; Grebenkov, 2007). These cell-type-specific assignments underpin the biological interpretation of our DW-MRS findings in the tumour context.

In our study, the reduction in tNAA ADC in IDH-wildtype tumours aligns with the known loss of neuronal integrity in higher-grade gliomas. tNAA is a well-established neuronal marker (Gill et al., 1989; Simmons et al., 1991; Petroff et al., 1993), and a decrease in its diffusion coefficient is consistent with displacement or restriction of intracellular diffusion associated with neuronal loss or architectural distortion such as cell swelling. The relative stability of tCho ADC across IDH mutation status may reflect the ubiquity of glial reaction irrespective of IDH mutation status, or may indicate that the diffusion properties of the glial compartment are less directly linked to tumour aggressiveness.

The VERDICT findings build on previous work demonstrating the utility of multi-compartment diffusion modelling for tumour characterisation. (Roberts et al., 2020; Figini et al., 2023) The significant reduction in extracellular diffusivities (D and D) in both enhancing and non-enhancing areas of IDH-wildtype lesions is noteworthy and may reflect increased density and remodelling of the ECM associated with the invasive growth pattern characteristic of GBM (Lan et al., 2024). This finding is particularly informative when considered alongside the ADC data: whereas standard ADC shows a reduction in enhancing and a non-significant trend towards elevation in non-enhancing areas (potentially reflecting co-existing oedema), VERDICT, leveraging its multi-compartment design tailored on tumour tissue components, is able to disentangle free-water effects from extracellular hindered diffusion, providing a more specific and interpretable microstructural signature of tumour infiltration.

The correlations between tCho ADC and VERDICT parameters in non-enhancing tumour are conceptually consistent, as both metrics reflect properties of the non-necrotic, infiltrative component of the tumour. tCho is enriched in glial cells (Choi et al., 2007), and its diffusion properties within the tumour may covary with the extracellular milieu, including ECM composition and free-water content, that VERDICT captures independently from the water signal. These preliminary observations motivate formal hypothesis-driven investigation in larger cohorts.

The combined DW-MRI and DW-MRS approach is technically demanding. Implementing cardiac-triggered DW-MRS on a clinical platform required a research agreement, bespoke pulse sequence programming and on-site support for modifications. Spectral data quality within the tumour was lower than in contralateral NAWM, possibly due to unsuccessful water suppression and areas of tumour necrosis, leading to a notable rate of data exclusion. Optimisation of the DW-MRS acquisition, including voxel positioning strategy, motion mitigation, and spectral quality thresholds aligned with current consensus recommendations (Ligneul et al., 2024), will be essential for future multicentre studies.

Several limitations of this study must be acknowledged. The sample size is small (n = 14 for DW-MRI; n = 10 for DW-MRS), and the cohort is heterogeneous in terms of WHO grade, IDH status, and treatment history at imaging. Statistical comparisons therefore have limited power, and results should be interpreted as hypothesis-generating. Furthermore, histopathological ground truth at the voxel level was unavailable, precluding direct validation of imaging-derived microstructural estimates. Future work should incorporate co-registered stereotactic biopsy or ex vivo correlative histology to establish the biological correspondence of VERDICT and DW-MRS biomarkers.

In conclusion, this study provides proof-of-concept that DW-MRI and DW-MRS can be combined within a clinically feasible protocol to probe complementary aspects of glioma cells and the glioma TME. VERDICT-MRI captured cell-level and ECM-level differences between IDH mutation strata, while DW-MRS provided metabolite-specific measures of neuronal and glial compartment integrity. Larger studies with histopathological validation are warranted to determine whether these combined biomarkers offer added diagnostic or prognostic value beyond standard imaging.

## Data Availability

All data produced in the present study are available upon reasonable request to the authors.

## Acknowledgements

The authors thank the patients who participated in this study and the MRI radiographers at UCL for their assistance with data acquisition

---

**Supplementary Table 1:**
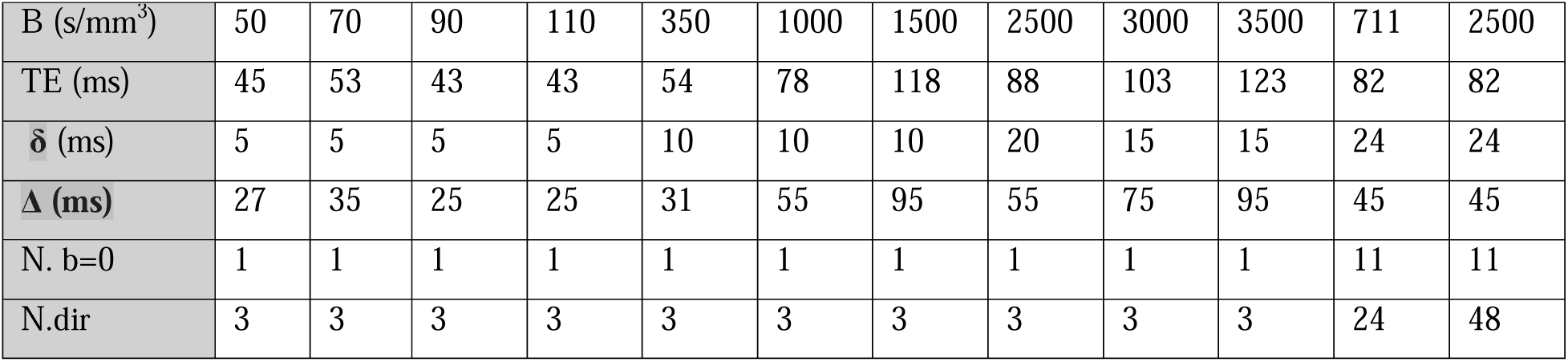
DW-MRI acquisition parameters.

